# Exploring patient-, provider-, and health facility-level determinants of blood pressure among patients with hypertension: A multicenter study in Ghana

**DOI:** 10.1101/2023.06.08.23291180

**Authors:** Samuel Byiringiro, Thomas Hinneh, Yvonne Commodore-Mensah, Jill Masteller, Fred Stephen Sarfo, Nancy Perrin, Shadrack Assibey, Cheryl R. Himmelfarb

## Abstract

Optimal blood pressure (BP) control is essential in averting cardiovascular disease and associated complications. Multi-level factors influence the achievement of BP targets in hypertension management. We explored patient-, provider-, and health facility-level factors of systolic and diastolic BP and controlled BP status among patients with hypertension in Ghana, where the burden of hypertension and cardiovascular disease is rising. Using a cross-sectional design, we recruited 15 health facilities, and from each facility, four healthcare professionals who managed patients with hypertension and 15 patients diagnosed with hypertension. The primary outcome of interest was systolic and diastolic BP; the secondary outcome was BP control (<140/90 mmHg) in compliance with Ghana’s national standard treatment guidelines. We used mixed-effects regression models to explore the patient- and facility-level predictors of the outcomes. From the 15 health facilities, we recruited 67 healthcare providers with a mean (SD) age of 32 (7) and 224 patients with a mean (SD) age of 60.5 (12.7). Most (182 [81%]) of the patient participants were female, and almost half (109 [49%]) had controlled BP. At the patient level, secondary school education (Coeff.:-7.69, 95% CI: -15.13, -0.26) was associated with lower systolic BP than a lower level of education; and traveling for 30 minutes to 1 hour to the health facility was associated with higher diastolic BP (Coeff.:3.75, 95% CI: 0.12, 7.38) and lower odds of BP control (OR: 0.51, 95% CI: 0.28, 0.92) than traveling less than 30 minutes. Receiving care at government health facilities was associated with less systolic BP (Coeff.: - 19.4; 95% CI: -33.58, -5.22) than private health facilities. A higher patient-to-physician or physician assistant ratio was associated with more elevated systolic BP (Coeff.: 23.06, 95% CI: 10.06, 36.05) and lower odds of controlled BP (OR: 0.18, 95% CI: 0.05, 0.74). Further research is warranted to identify effective strategies to address the multi-level factors influencing hypertension control and to mitigate the rising burden of hypertension in Ghana.

## Background

A quarter of the adult population in Sub-Saharan Africa (SSA) is estimated to be living with chronic hypertension, a major risk factor for leading causes of death, cardiovascular disease, and stroke [1,2]. In Ghana, approximately 27% of the adult population has hypertension, yet 65% of the patients are unaware of their hypertension status [3]. Twenty-one percent of patients are on treatment, and only 27% of patients on treatment achieve controlled Blood Pressure (BP) (6% of all hypertensive patients)— which highlights the dire need for improving the quality of hypertension care services [3]. Countries in SSA are predominantly classified by the World Bank as low-income and are ill-equipped with frail health systems that are not prepared to respond to the projected further rise in hypertension and other non-communicable diseases (NCDs) [4].

Inadequate health systems financing contributes to the shortcomings across key health system components including the lack of essential equipment, proper training of staff, and the continuous stocking of health facility pharmacies with necessary consumables [5,6]. These shortcomings further limit the public health systems’ initiatives to raise the population’s awareness of the dangers of major NCDs such as hypertension, hence leaving most patients undiagnosed and therefore without access to treatment. Effective strategies are needed to respond to the rising burden of hypertension.

While hypertension is a chronic condition with life-threatening complications, these can be averted by controlling BP using lifestyle modifications, antihypertensive therapy, or both [7]. Multiple guidelines with different thresholds for defining hypertension and initiating patients on treatment exist [7–10]. The 2017 American Heart Association/American College of Cardiology guidelines recommend a treatment target of ≤130/80 mmHg [7] while the World Heart Federation in its 2021 roadmap for hypertension management as well as many countries in SSA recommend a treatment to target of ≤ 140/90 mmHg [10]. Despite variabilities in BP treatment targets, evidence shows that even among patients with uncontrolled hypertension, a 5-mmHg reduction in systolic or diastolic BP is associated with better health outcomes through the reduction of composite cardiovascular event risk [11,12]. It would hence be beneficial to explore factors not only of hypertension control, but also for systolic and diastolic BP reduction.

The factors that influence hypertension care and outcomes are multifaceted and exist at the individual, community, and health system levels. The individual-level factors that have shown an association with hypertension include age, sex, socioeconomic status, education, employment, family history of hypertension, and lifestyles such as smoking, high salt consumption, alcoholism, and the lack of physical exercise [13]. At the community level, the lack of resources (or policy to ensure the availability of the resources) such as patient peer support groups, non- access to the facilities for physical exercises, and affordable foods for healthy diets contribute to the worsening status of hypertensive patients [14,15]. There are very interlinked influences among the various risk levels of acquiring or having a worsened state of hypertension. Without strong health facilities and systems, the population sensitization and screening for elevated BP are very weak. At the health facilities, provider factors such as inadequate training and shortages of healthcare providers could contribute to poor BP outcomes among patients already on hypertension treatment. These factors make the exploration of multi-level factors of hypertension care and outcomes very important.

Health facilities play a critical role in coordinating screening for raised BP and ensuring that identified patients achieve desired BP targets. The World Health Organization (WHO)’s health systems framework highlights six key factors of health systems strengthening to look out for including: service delivery, health workforce, medications, diagnostics, equipment, and technology, health information systems, financing, and leadership and governance [16]. In the context of hypertension care, health facilities require a sufficient number of well-trained healthcare providers, validated and regularly inspected BP devices, and other essential tools for taking physical measurements, sufficient stock of evidence-based anti-hypertensive medications, and all the systems for ensuring that diagnosed patients are initiated and maintained on treatment. The aim of this study was to explore the health facility readiness for hypertension management and assess the patient-, provider- and health facility-level factors of systolic and diastolic BP and BP control among patients receiving hypertension care at 15 health facilities in Kumasi, Ghana.

## Methods

### Setting

We conducted this study in the Kumasi Metropolitan District, the capital city of the Ashanti region in Ghana [17]. According to the 2010 population and housing census in Ghana, the Kumasi Metropolitan District had a population of 1,730,249 (36% of the Ashanti region’s population) but grew up to approximately 2,096,053 people by 2019 [18,19]. The Kumasi Metropolitan District is served by 263 health facilities, 145 of which are clinics, three health centers, 53 hospitals, six training institutions, 55 maternity homes, and one regional hospital. Of all the health facilities, 224 are privately owned, 18 are quasi- governmental, 16 are government owned, and five are owned by the Christian Health Association of Ghana (CHAG) [20].

### Design

We used a cross-sectional study design to explore the patient-, provider-, and health facility-level determinants of systolic and diastolic BP and the overall BP control. We followed the Strengthening the Reporting of Observational Studies in Epidemiology (STROBE) checklist [21] to ensure an elaborate and unbiased reporting and discussion of the findings (S1 Table).

### Participants

We used a convenience sample of 15 health facilities at different levels of the health system and ownership. Eligible health facilities were as follows: (a) located in Kumasi Metropolitan District; (b) provided hypertension service per the ministry of health structure; (c) qualified as government, quasi-governmental, private or Christian Health Association of Ghana hospital, regional or tertiary hospitals, Clinics, or community health centers; and (d) had leadership willing to be part of the study. From each participating health facility, we recruited four healthcare providers involved in managing patients with hypertension. All clinical care providers—including nurses, physicians, physician assistants, and dietitians working in hypertension care clinics or outpatient departments (OPD)—were eligible to participate in the study.

Additionally, from each health facility, we recruited 15 patients diagnosed with hypertension and prescribed antihypertensive therapy. We included patients 18 years of age and older.

We conducted the recruitment and data collection between April 15^th^ and June 1^st^, 2022.

### Recruitment and data collection process

We traveled to the health facilities for data collection after receiving the signed memorandum of understanding from the health facilities. We dedicated a minimum of two days per health facility for data collection. After arriving at the health facility, a pair of trained enumerators, led by the ranking health facility leader, visited the different health facility units to collect facility-level data about hypertension. Another enumerator recruited potential participants among healthcare providers to complete the self-administered survey at that time or take the link and complete the survey later. On the second day, the team recruited patients coming for their usual hypertension care appointments. Before data collection, both the providers and patients signed the consent form, electronically for healthcare providers and on paper for patients,. We compensated patients and providers with 10 USD and 25 USD, respectively, for their time.

Outcome of interest

The primary outcome was the patients’ systolic and diastolic BP. We treated the systolic and diastolic BP as continuous variables. To measure patients’ BP, we used Omron 7 Series Upper Arm BP Monitor following the American Heart Association’s guideline for BP measurement [22]. After 3 minutes of patient rest, we took three consecutive measures of BP with 1 minute between each measurement and used the average of the last two for BP measurement ascertainment.

### Secondary outcomes

The secondary outcome was BP control. We adopted the treatment goal stated by the Ghana hypertension treatment guidelines to define BP control (<140/90 mmHg) [23]. We hence treated the BP control as a binary variable.

### Exposures of interest

The exposures of interest were at the health facility and patient level.

#### Patient level exposures

We measured the patient behaviors regarding adherence to high BP treatment using the Hill-Bone Compliance to High Blood Pressure Therapy Scale (HB-HBP), a 14-item scale with an internal consistency reliability of 0.85 [24]. HB-HBP has 4 choice options on the Likert scale ranging from “All of the Time” to “None of the Time” valid for 1 to 4 points respectively in that order except for item 6 “How often do you make the next appointment before you leave the doctor’s office?” which requires reverse coding. HB-HBP scores ranged from 14 to 56, with higher scores indicating behaviors of higher adherence to high BP therapy. HB-HBP consists of three sub-scales: (1) dietary sodium intake, three items with score ranging from 3 to12; (2) medication adherence, nine items with score ranging from 9 to 36; and (3) appointment keeping, two items with score ranging from 2 to 8. We treated the HB-HBP overall and subscale scores as continuous variables.

The other self-reported exposure variables that we measured at the patient level were the length of time with hypertension diagnosis, which we treated as a binary variable (<five years, ≥five years) and travel duration to the health facility where they received hypertension care services as a categorical variable (< 30 minutes, 30 minutes to < one hour, and > one hour).

#### Healthcare provider level exposures

The provider-level predictors were the provider knowledge of and attitude towards hypertension treatment guidelines. In Ghana, patients are not assigned a primary care provider who would ideally follow them up over time. They rather usually meet the healthcare provider who is in the consulting office on the day of their appointment. For this reason, we could not link specific provider responses with a given patients’ BP outcomes. We hence could not ascertain the association between patient BP and the provider knowledge score or attitudes towards hypertension guidelines.

We measured provider knowledge of hypertension treatment guidelines using a 23-item questionnaire developed by the investigation team. Five items were multiple choice questions with four response options, six were multiple choice questions with three response options plus “I don’t know”, and 14 items were true and false questions with “I don’t know” as added response option. Each of the 23 items was scored at one point, and assigned if the provider made the correct selection. We reported the total scores out of 23, with higher scores indicating higher knowledge of hypertension guidelines.

To measure the provider attitude towards hypertension guidelines, we assessed their self-reported confidence in adhering to hypertension treatment guidelines. To measure self-reported confidence, we asked them to rate on a scale of “Very confident” to “Not at all confident” their adherence to guidelines while (1) measuring BP, (2) assigning hypertension diagnosis, (3) educating patients with hypertension on healthy lifestyles, (4) prescribing antihypertensive medications, and (5) their overall management of patients with hypertension. In addition to this 5-point Likert scale, we had a “not applicable” option for the tasks that were outside the provider’s scope of practice.

#### Health facility level exposures

We used the WHO Service Availability and Readiness Assessment Tool to collect health facility-level measurements of hypertension service readiness [25]. Health facility characteristics included bed capacity as a categorical variable (<50, 50 to <100 beds, and ≥100 beds), facility ownership as a categorical variable (government, private or Christian Health Association of Ghana (CHAG) health facilities), and possession of a hypertension clinic (versus managing patients in the general OPD) as a binary variable.

The indicators related to health workforce that we measured were the nurse-to-patient ratio and physician and physician assistant-to-patient ratio. To calculate the clinician-to-patient ratio, we divided the number of patients received in the OPD in one recent month with the number of registered and enrolled nurses or the number of physicians (generalist and specialists) and physician assistants. We combined the physician and physician assistants because they were licensed to diagnose and prescribe anti-hypertensive medications in their scope of practice in Ghana. We categorized the patient-to-nurse ratio as < 20, 20 to < 40, and ≥ 40 patients per nurse per month and the patient-to-physician or physician assistant ratio <140, 140 to < 280, and ≥ 280 patients per physician (or assistant) per month. We additionally asked if at least one healthcare provider had received training on hypertension treatment guidelines over the previous two years and treated this indicator as a binary variable.

The hypertension service readiness indicators we assessed were the availability of the 2017 Ghana Standard treatment guidelines [26]or the 2019 National Guidelines for the Management of Cardiovascular Diseases (first edition) [23]; patient education materials consisting of brochures and risk charts; BP measurement apparatus (functional automated BP device or a sphygmomanometer with a stethoscope); the basic laboratory exams essential for patients with hypertension, and all first line antihypertensive medications according to Ghana protocol [27].

The basic laboratory exams included were as follows: (1) valid dipstick for urine protein, (2) urine glucose, and (3) urine ketones, (4) glucometer for blood glucose and valid test strips, and (5) hemoglobinometer or analyzer for hemoglobin testing. The Ghana the Standard Treatment Guidelines 7^th^ Edition [26] and the Essential Medicines List 7^th^ Edition [27] require each health facility that treats hypertension to possess all the first line antihypertension medications: Angiotensin Converting Enzyme Inhibitors, Angiotensin Receptor Blocker, Calcium Channel Blocker, Thiazide, and Beta Blocker.

#### Covariates

The patient-level covariates we explored included patient age, educational level, employment status, and marital status. We treated patient age as a continuous variable; biological sex as a binary variable (male, female); education as a categorical variable (primary education or less, secondary education, and tertiary education or higher); employment status as a categorical variable (unemployed, employed, and retired); and marital status as a binary variable (married or cohabiting and single, separated or widowed).

The provider-level covariates we evaluated were age, sex, educational level, profession, and experience. We treated provider age as a continuous variable and sex as a binary variable (male, female), education as a categorical variable (associate degree or less, bachelor’s degree, and post-graduate degree), and profession as a categorical variable (physicians (generalist, specialist, physician assistant), and nurses (enrolled and registered), and other clinical staff). We treated experience as a categorical variable (<two years, two to <four years, ≥four years).

We additionally asked providers whether they had received hypertension management training over the previous two years (yes or no) and the type of training they had received.

### Analysis

We explored patients’ characteristics by BP control status. We used means (±SD) to report continuous variables and proportions and percentages for categorical variables. We used two sample *t*-tests to compare means of the continuous variables and chi-square tests to compare the proportions of the categorical variables by the BP control status.

We used percentages to summarize provider characteristics and means to summarize provider scores of hypertension guidelines knowledge. To compare scores of hypertension guidelines knowledge across the different categorical variables of provider characteristics, we used one-way analysis of variance. For the provider attitude towards hypertension guidelines, we reported percentages of providers in each level of self-reported confidence in adherence to the guidelines. We used a stacked graph to report the provider’s levels of self confidence in adhering to hypertension treatment guidelines. Since the health facility-level data were limited in the number of observations, we reported proportions (proportion of health facilities which met a certain standard of hypertension care) and percentages for categorical variables, and the median and interquartile ranges (IQR) for continuous variables.

We used mixed-effects linear regression models to explore the patient- and health facility-level factors of systolic and diastolic BP, and used mixed-effects logistic regression models to explore the patient- and health facility-level factors of BP control. We chose this model to account for possible intra-cluster correlation in BP outcomes across the different health facilities. We initially estimated the bivariate models of each patient and facility level predictor then fitted fully adjusted models separately for patient- and facility-level predictors. At the patient-level, the fully adjusted models included the hypertension adherence variables, travel distance to the health facility, duration with hypertension diagnosis, and all sociodemographic characteristics listed under covariates. At the facility level, the fully adjusted model included the health facility characteristics (ownership, availability of hypertension clinic, bed capacity); health workforce indicators (patient-to-clinician ratio and provider training); and hypertension service availability indicators (availability of all first line antihypertensive medications). The distribution of data for these health facility level variables—basic laboratory exams, BP devices, and hypertension guidelines; and patient educational materials—rendered the fully adjusted model unstable and were excluded. We reported the coefficients or odds ratio with the associated 95% confidence intervals. We considered associations with 0.05 or smaller *p*-values to be statistically significant.

The power analysis assumed an Intra-Cluster Correlation (ICC) of 0.1 based on the systolic BP ICC reported by the Phone-based Intervention under Nurse Guidance after Stroke (PINGS) study interim results [28] and a BP control rate of 27% among hypertensive patients on treatment [3]. With these assumptions, our mixed-effects logistic regression had 80% power to predict odds ratios of BP control of 1.92 or greater with continuous predictor variables with alpha of 0.05. For dichotomous predictor variables, odds ratios of 3.31, 3.37, and 4.28 were to be detectable if the predictor variable has a 50/50, 40/60, and 20/80 split, respectively.

## Results

### Patient Characteristics

This study included 224 patients with a mean (SD) age of 60.5 (12.7). Most participants, 182 (81%) were female. The mean (SD) systolic BP was 139 (20) mmHg and diastolic BP was 85 (13) mmHg (Table 1). Almost half of the participants, 109 (49%) participants had controlled BP (<140/90 mmHg). A little over half of the patients (54%) were single, separated or widowed, almost half of the patients (47%) were employed and 199 (45%) of the patients had a tertiary or higher level of education. One hundred and forty-two (64%) patients had lived with a hypertension diagnosis for more than 5 years.

**Table 1.**
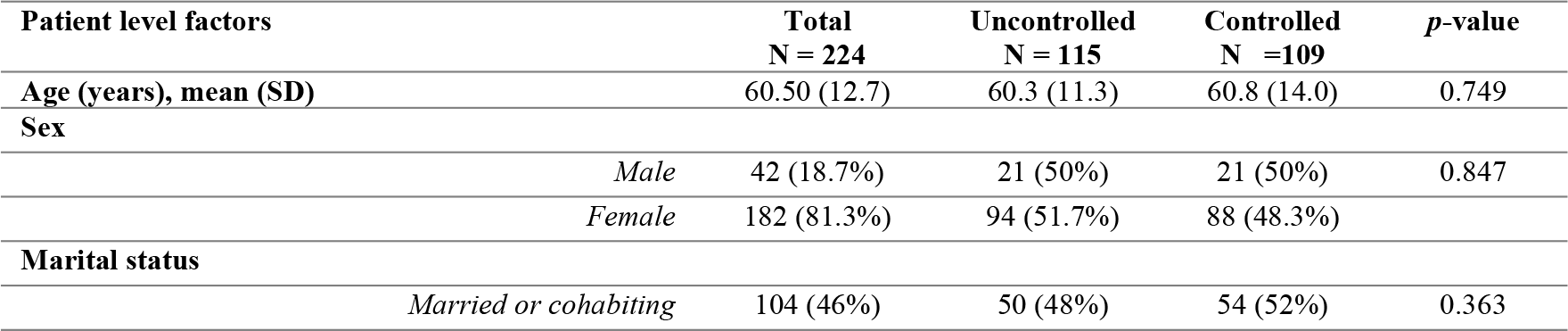
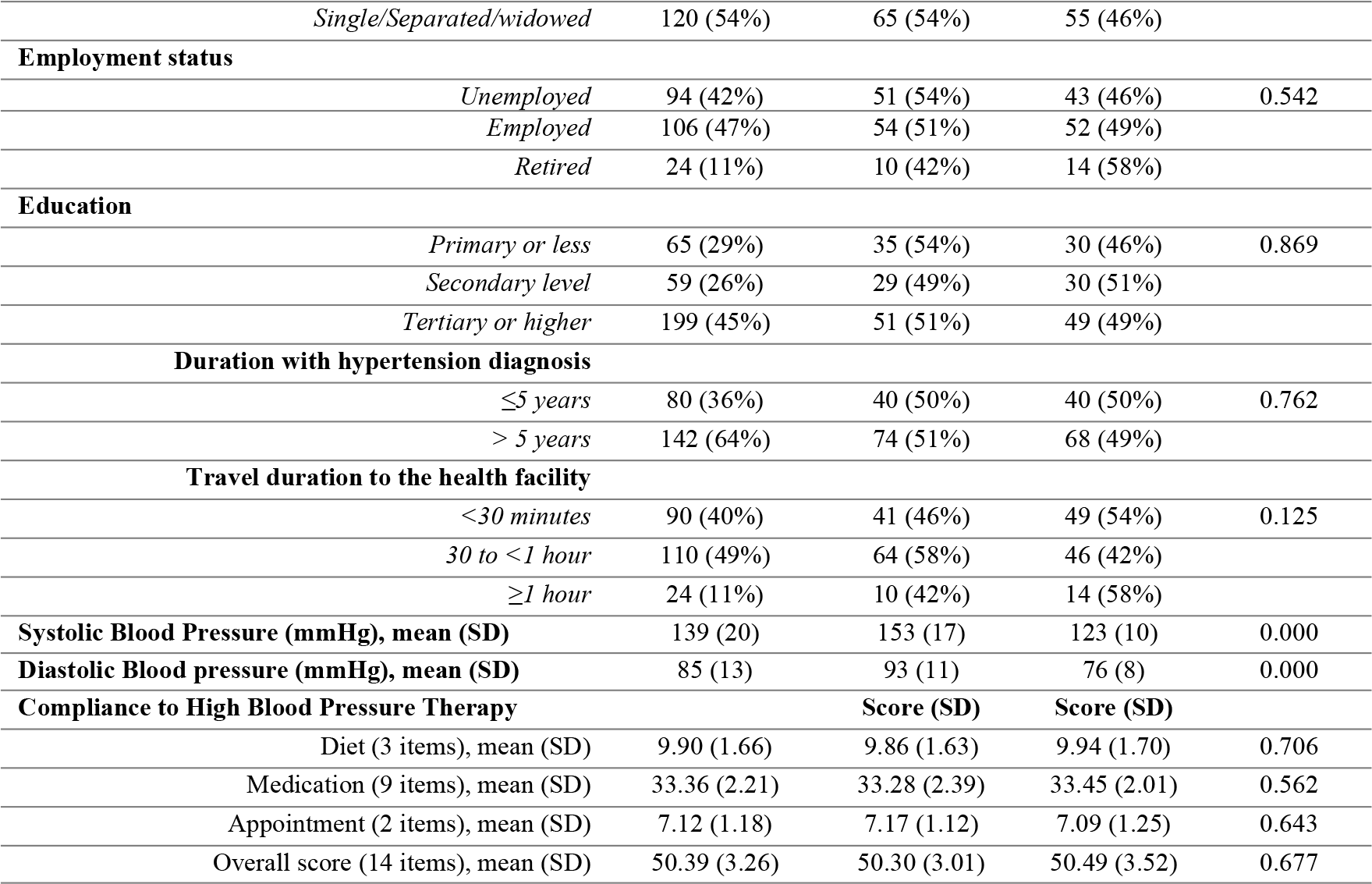
Patients Characteristics stratified by Blood Pressure Control Status (BP <140/90 mmHg) Status, n (%) or mean (SD) (N = 224)

| Patient level factors | Total<br>N = 224 | Uncontrolled<br>N = 115 | Controlled<br>N = 109 | $p$-value |
| --- | --- | --- | --- | --- |
| Age (years), mean (SD) | 60.50 (12.7) | 60.3 (11.3) | 60.8 (14.0) | 0.749 |
| Sex |  |  |  |  |
| Male | 42 (18.7%) | 21 (50%) | 21 (50%) | 0.847 |
| Female | 182 (81.3%) | 94 (51.7%) | 88 (48.3%) |  |
| Marital status |  |  |  |  |
| Married or cohabiting | 104 (46%) | 50 (48%) | 54 (52%) | 0.363 |

|  |  |  |  |  |  |
| --- | --- | --- | --- | --- | --- |
|  | *Single/Separated/widowed* | 120 (54%) | 65 (54%) | 55 (46%) |  |
| **Employment status** |  |  |  |  |  |
|  | *Unemployed* | 94 (42%) | 51 (54%) | 43 (46%) | 0.542 |
|  | *Employed* | 106 (47%) | 54 (51%) | 52 (49%) |  |
|  | *Retired* | 24 (11%) | 10 (42%) | 14 (58%) |  |
| **Education** |  |  |  |  |  |
|  | *Primary or less* | 65 (29%) | 35 (54%) | 30 (46%) | 0.869 |
|  | *Secondary level* | 59 (26%) | 29 (49%) | 30 (51%) |  |
|  | *Tertiary or higher* | 199 (45%) | 51 (51%) | 49 (49%) |  |
| **Duration with hypertension diagnosis** |  |  |  |  |  |
|  | *≤5 years* | 80 (36%) | 40 (50%) | 40 (50%) | 0.762 |
|  | *> 5 years* | 142 (64%) | 74 (51%) | 68 (49%) |  |
| **Travel duration to the health facility** |  |  |  |  |  |
|  | *<30 minutes* | 90 (40%) | 41 (46%) | 49 (54%) | 0.125 |
|  | *30 to <1 hour* | 110 (49%) | 64 (58%) | 46 (42%) |  |
|  | *≥1 hour* | 24 (11%) | 10 (42%) | 14 (58%) |  |
| **Systolic Blood Pressure (mmHg), mean (SD)** |  | 139 (20) | 153 (17) | 123 (10) | 0.000 |
| **Diastolic Blood pressure (mmHg), mean (SD)** |  | 85 (13) | 93 (11) | 76 (8) | 0.000 |
| **Compliance to High Blood Pressure Therapy** |  |  | **Score (SD)** | **Score (SD)** |  |
|  | Diet (3 items), mean (SD) | 9.90 (1.66) | 9.86 (1.63) | 9.94 (1.70) | 0.706 |
|  | Medication (9 items), mean (SD) | 33.36 (2.21) | 33.28 (2.39) | 33.45 (2.01) | 0.562 |
|  | Appointment (2 items), mean (SD) | 7.12 (1.18) | 7.17 (1.12) | 7.09 (1.25) | 0.643 |
|  | Overall score (14 items), mean (SD) | 50.39 (3.26) | 50.30 (3.01) | 50.49 (3.52) | 0.677 |

The HB-HBP Scale mean scores (+SD) were: Overall, 50.39 (+3.26); medication adherence, 33.36 (+2.21); appointment keeping, 7.12 (+1.18); and dietary sodium intake, 9.90 (+1.66).

There was no statically significant difference in binary comparison by BP control status with any of the measured variables

### Provider Characteristics

Healthcare providers (N = 67) were 32 (+7) years of age (Table 2). Most healthcare providers, 53 (79%) were female, 40 (61%) had associate degree or less, and the majority, 61 (91%) were enrolled or registered nurses. Of the healthcare providers, 32 (48%) had 4 or more years of experience, 39 (59%) worked in general OPD, and 44 (66%) reported that they had received training on hypertension management in the previous 2 years.

**Table 2.** Healthcare professional characteristics (n = 67)

| Characteristic | Number (%) |
| --- | --- |
| Mean Hypertension Knowledge Score (SD) | 18.9 (1.9) |
| **Age in years, mean (SD)** | 32 (7) |
| **Sex** |  |
| *Male* | 14 (21%) |
| *Female* | 53 (79%) |
| **Education (n = 66)** |  |
| *Associate degree or less* | 40 (61%) |
| *Bachelor's degree* | 20 (30%) |
| *Post-graduate degree* | 6 (9%) |
| **Profession** |  |
| *Physician/Doctors* | 6 (9%) |
| *Enrolled & Registered Nurses* | 55 (82%) |
| *Other clinical care providers* | 6 (9%) |
| **Experience** |  |
| *< 2 years* | 19 (28%) |
| *2 to <4 years* | 16 (24%) |
| *≥4 years* | 32 (48%) |
| **Health facility department** |  |
| *Hypertension Clinic* | 10 (15%) |
| *General OPD* | 39 (59%) |
| *Others* | 17 (26%) |
| **Received hypertension training in the last 2 years** |  |
| *No* | 23 (34%) |
| *Yes* | 44 (66%) |
| **Type of hypertension treatment received (n = 44)** |  |
| *Job orientation* | 13 (30%) |
| *On job training* | 15 (34%) |
| *Professional certification program* | 7 (16%) |
| *Conference or other hypertension training workshops* | 9 (20%) |

### Health facility Characteristics

We conducted the study at 15 health facilities, among which almost half (47%) had less than 50- bed capacity, and nearly half (47%) were government-owned (Table 3 and Table 4). Eight (53%) of the health facilities managed patients in hypertension clinic; the remaining seven managed them in the general OPD.

**Table 3.** Characteristics of included health facilities in Ashanti Region, Ghana (N = 15)

| **Facility characteristics, n (%) or median (IQR)** | **Sites** |
| --- | --- |
| **Facility size (by number of beds)** |  |
| *<50 beds* | 7 (47%) |
| *50 to <100 beds* | 5 (33%) |
| *≥100 beds* | 3 (20%) |
| **Facility ownership** |  |
| *Government* | 7 (47%) |
| *Private* | 5 (33%) |
| *Christian Health Association of Ghana* | 3 (20%) |
| **Patients treated in hypertension clinic** |  |
| *No (General OPD only)* | 7 (47%) |
| *Yes* | 8 (53%) |
| **Personnel and training on cardiovascular diseases' management** |  |
| Number of full-time nurses (registered and enrolled), median (IQR) | 35 (24-205) |
| Number of full-time doctors (generalists and specialists), median (IQR) | 9 (5-17) |
| Number of full-time paramedical clinicians, median (IQR) | 2 (1-7) |
| Patient/nurse ratio, media (IQR) – based on patient received in OPD last month | 26 (19-46) |
| Patient/physician ratio, media (IQR) – based on patient received in OPD last month | 235 (139-388) |
| Number of facilities where at least one provider received training on cardiovascular diseases within the last 2 years, n (%) | 11 (73%) |
| **Hypertension Service Readiness Indicators, n (%)** |  |
| Hypertension guidelines<sup>0</sup> | 12 (80%) |
| Patient education materials | 9 (60%) |
| At least one functional blood pressure measurement apparatus<sup>1</sup> | 15 (100%) |
| Basic laboratory exams available<sup>2</sup> | 11 (73%) |
| All first line treatment medications available (n = 14)<sup>3</sup> | 9 (64%) |
| At least one second line treatment medication available (n = 14)<sup>4</sup> | 14 (100%) |
| **Availability of Essential antihypertensive medications, n = 14 (%)** |  |
| Angiotensin converting enzyme inhibitor | 14 (100%) |
| Angiotensin receptor blocker | 13 (93%) |
| Calcium channel blocker | 14 (100%) |
| Thiazide diuretic | 9 (64%) |
| Beta blocker | 11 (78%) |
| Aldosterone antagonist | 4 (28%) |
| Central acting agent | 12 (86%) |
| Vasodilator | 10 (71%) |
| Fixed dose combination | 8 (57%) |

Abbreviations: IQR – Interquartile Range; OPD – Outpatient Department.
<sup>0</sup>Considered the 2017 national treatment standards or 2021 cardiovascular diseases guidelines.
<sup>1</sup>Considered automatic Blood Pressure device or sphygmomanometer with stethoscope.
<sup>2</sup>Included 5 tests: valid dipstick for urine protein, urine glucose, and urine ketones; glucometer for blood glucose and valid test strips; and hemoglobinometer or analyzer for hemoglobin testing.
<sup>3</sup>First line treatment medications considered according to Ghana Protocol - Angiotensin Converting Enzyme Inhibitors, Angiotensin Receptor Blocker, Calcium Channel Blocker, Thiazide, and Beta Blocker.
<sup>4</sup>Second line treatment medications considered - Centrally Acting Agent; Vasodilators, and Aldosterone Antagonists.

**Table 4.** Health Facility Characteristics Grouped by Patient Blood Pressure Control Status of Included Patients, n (%) (N = 224)

| Patient level factors | Total<br>N=224 | Uncontrolled<br>N=115 | Controlled<br>N =109 | p-value |
| --- | --- | --- | --- | --- |
| Facility ownership |  |  |  |  |
| Private | 69 (31%) | 33 (48%) | 36 (52%) | 0.659 |
| Government | 109 (49%) | 56 (51%) | 53 (49%) |  |
| CHAG | 46 (20%) | 26 (57%) | 20 (43%) |  |
| Hypertension Clinic |  |  |  |  |
| General OPD only | 101 (45%) | 57 (56%) | 44 (44%) | 0.167 |
| Yes | 123 (55%) | 58 (57%) | 65 (53%) |  |
| Provider training |  |  |  |  |
| No | 53 (24%) | 25 (47%) | 28 (53%) | 0.487 |
| Yes | 171 (76%) | 90 (53%) | 81 (47%) |  |
| Facility size by bed count |  |  |  |  |
| < 50 beds | 100 (45%) | 50 (50%) | 50 (50%) | 0.682 |
| 50 to < 100 beds | 76 (34%) | 42 (55%) | 34 (45%) |  |
| ≥ 100 beds | 48 (21%) | 23 (48%) | 25 (52%) |  |
| **All first line medications available** |  |  |  |  |
| No | 67 (32%) | 33 (49%) | 34 (51%) | 0.666 |
| Yes | 143 (68%) | 75 (52%) | 68 (48%) |  |
| **Patient to nurse ratio** (patients per month per nurse) |  |  |  |  |
| < 20 | 76 (30%) | 31 (46%) | 36 (54%) | 0.611 |
| 20 to <40 | 99 (44%) | 53 (54%) | 46 (46%) |  |
| ≥40 | 58 (26%) | 31 (53%) | 27 (47%) |  |
| **Patient to physician or physician assistant ratio** (patients per month per clinician) |  |  |  |  |
| < 140 | 74 (33%) | 31 (42%) | 43 (58%) | 0.095 |
| 140 to <280 | 90 (40%) | 53 (59%) | 37 (41%) |  |
| ≥280 | 60 (27%) | 31 (52%) | 29 (48%) |  |

Regarding the health facilities’ hypertension service readiness indicators, 11 (73%) health facilities had at least one healthcare provider who received training on cardiovascular diseases within the last 2 years. Hypertension guidelines were available in 12 (80%) health facilities, patient education materials were available in nine (60%) health facilities, and all 15 (100%) health facilities had at least one functional BP measurement apparatus. The basic laboratory exams were available in 11 (73%) health facilities, and all first-line antihypertensive medications were available in nine (64%) of 14 health facilities assessed for medications availability. All 14 (100%) health facilities had at least one second-line antihypertensive medication. Angiotensin- converting enzyme inhibitors were available in all 14 (100%) health facilities, and thiazide diuretics were the least available (observed in 9 or 64% of the 14 assessed health facilities) of the first-line antihypertensive medications. Of the second-line antihypertensive medications, a centrally acting agent category of drugs was the most available (observed in 12 or 86% of the 14 assessed health facilities); aldosterone antagonist category was the least available (observed in 4 or 28% of the 14 assessed health facilities).

Patient-, provider-, and facility level predictors systolic and diastolic BP

#### Patient-level predictors of systolic and diastolic BP and BP control

The patients’ educational level was associated with systolic BP while travel duration to the health facility, and the overall HB-HBP scores were associated with diastolic BP. Having a secondary level of education was associated with roughly 8 mmHg lower systolic BP (Coefficient: -7.69; 95% CI: -15.13, -0.26) than having a primary or lower educational level (Table 5). Additionally, travelling for an hour or more to the health facility was associated with roughly 7 mmHg higher (Coefficient: 6.86; 95% CI: 0.92, 12.81) diastolic BP. Finally, a unit increase in overall HB-HBP scores was associated with a ½ mmHg lower (Coefficient: -0.55, 95% CI: -1.09, -0.01) diastolic BP. In the HB-HBP sub-scores, a unit increase in the diet control scores was associated with one unit lower (Coefficient: -1.11; 95% CI: -2.20, -0.02) diastolic BP in mmHg.

**Table 5.**
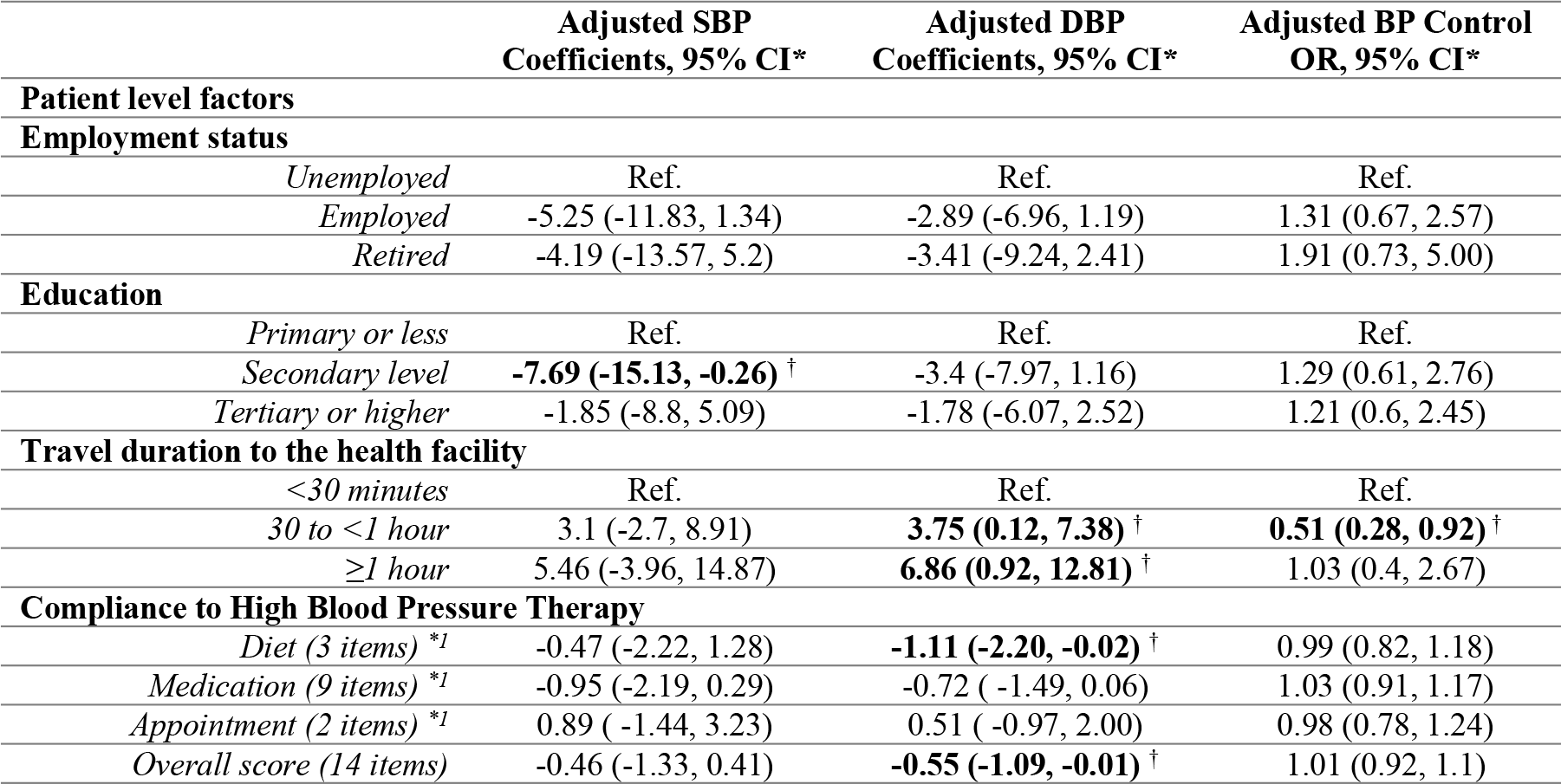
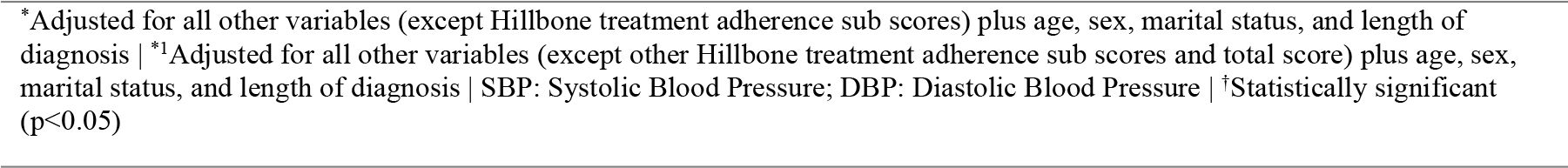
Mixed-Effects Regression of Patient Factors of Systolic and Diastolic Blood Pressure and Blood Pressure Control in Kumasi, Ghana (n = 221 patients, 15 health facilities)

|  | Adjusted SBP<br>Coefficients, 95% CI* | Adjusted DBP<br>Coefficients, 95% CI* | Adjusted BP Control<br>OR, 95% CI* |
| --- | --- | --- | --- |
| **Patient level factors** |  |  |  |
| **Employment status** |  |  |  |
| *Unemployed* | Ref. | Ref. | Ref. |
| *Employed* | -5.25 (-11.83, 1.34) | -2.89 (-6.96, 1.19) | 1.31 (0.67, 2.57) |
| *Retired* | -4.19 (-13.57, 5.2) | -3.41 (-9.24, 2.41) | 1.91 (0.73, 5.00) |
| **Education** |  |  |  |
| *Primary or less* | Ref. | Ref. | Ref. |
| *Secondary level* | **-7.69 (-15.13, -0.26) †** | -3.4 (-7.97, 1.16) | 1.29 (0.61, 2.76) |
| *Tertiary or higher* | -1.85 (-8.8, 5.09) | -1.78 (-6.07, 2.52) | 1.21 (0.6, 2.45) |
| **Travel duration to the health facility** |  |  |  |
| *<30 minutes* | Ref. | Ref. | Ref. |
| *30 to <1 hour* | 3.1 (-2.7, 8.91) | **3.75 (0.12, 7.38) †** | **0.51 (0.28, 0.92) †** |
| *≥1 hour* | 5.46 (-3.96, 14.87) | **6.86 (0.92, 12.81) †** | 1.03 (0.4, 2.67) |
| **Compliance to High Blood Pressure Therapy** |  |  |  |
| *Diet (3 items) \*1* | -0.47 (-2.22, 1.28) | **-1.11 (-2.20, -0.02) †** | 0.99 (0.82, 1.18) |
| *Medication (9 items) \*1* | -0.95 (-2.19, 0.29) | -0.72 (-1.49, 0.06) | 1.03 (0.91, 1.17) |
| *Appointment (2 items) \*1* | 0.89 (-1.44, 3.23) | 0.51 (-0.97, 2.00) | 0.98 (0.78, 1.24) |
| *Overall score (14 items)* | -0.46 (-1.33, 0.41) | **-0.55 (-1.09, -0.01) †** | 1.01 (0.92, 1.1) |

\*Adjusted for all other variables (except Hillbone treatment adherence sub scores) plus age, sex, marital status, and length of diagnosis | <sup>†</sup>Adjusted for all other variables (except other Hillbone treatment adherence sub scores and total score) plus age, sex, marital status, and length of diagnosis | SBP: Systolic Blood Pressure; DBP: Diastolic Blood Pressure | <sup>†</sup>Statistically significant (p<0.05)

In the unadjusted and fully adjusted mixed-effects logistic regression models, only the duration of travel to the health facility remained significant. Patients who travelled 30 minutes to an hour to the health facility were two times less likely to have controlled BP than their counterparts who travelled less than 30 minutes (OR: 0.51; 95% CI: 0.28, 0.92).

#### Provider level predictors of BP outcomes

Provider knowledge of hypertension guidelines was 18.9 (+1.9) on a 23-point scale. In association analyses, no provider characteristics were associated with the provider knowledge scores. Two-thirds (75% of the 64) of respondents reported being very or moderately familiar with hypertension treatment guidelines (Fig 1).

**Fig 1.**
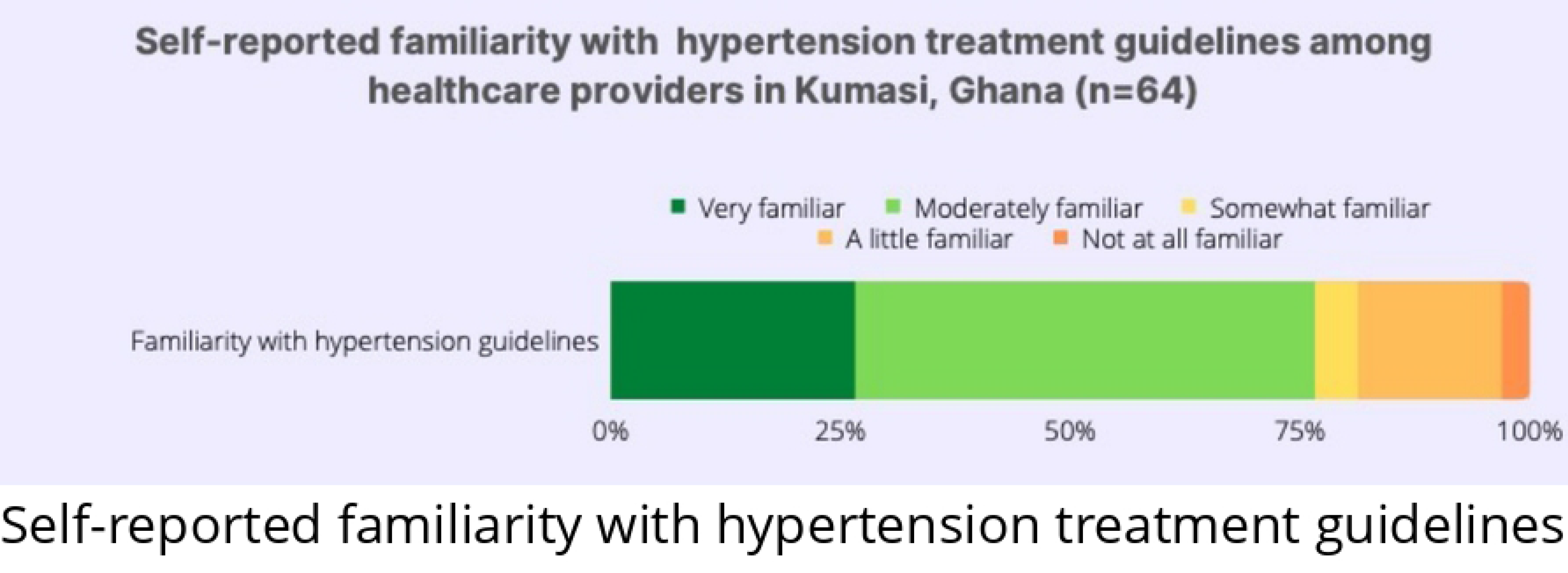
Self-reported familiarity with hypertension treatment guidelines among healthcare providers in Kumasi, Ghana (n = 64)

Regarding the nurses’ and other clinical staffs’ (other than medical doctors) attitudes towards hypertension guidelines, almost 100% of the respondents were “very confident” in their adherence to the guidelines while measuring BP; roughly 80% were “very confident” in providing counselling on healthy lifestyles (Fig 2). Only half of the respondents were “very confident” in diagnosing hypertension, and even fewer, roughly 30%, were confident in prescribing antihypertensive medications. Overall, 75% of the nurses and other clinical staff (except medical doctors) were very confident in adhering to hypertension guidelines while managing patients with hypertension.

**Fig 2.**
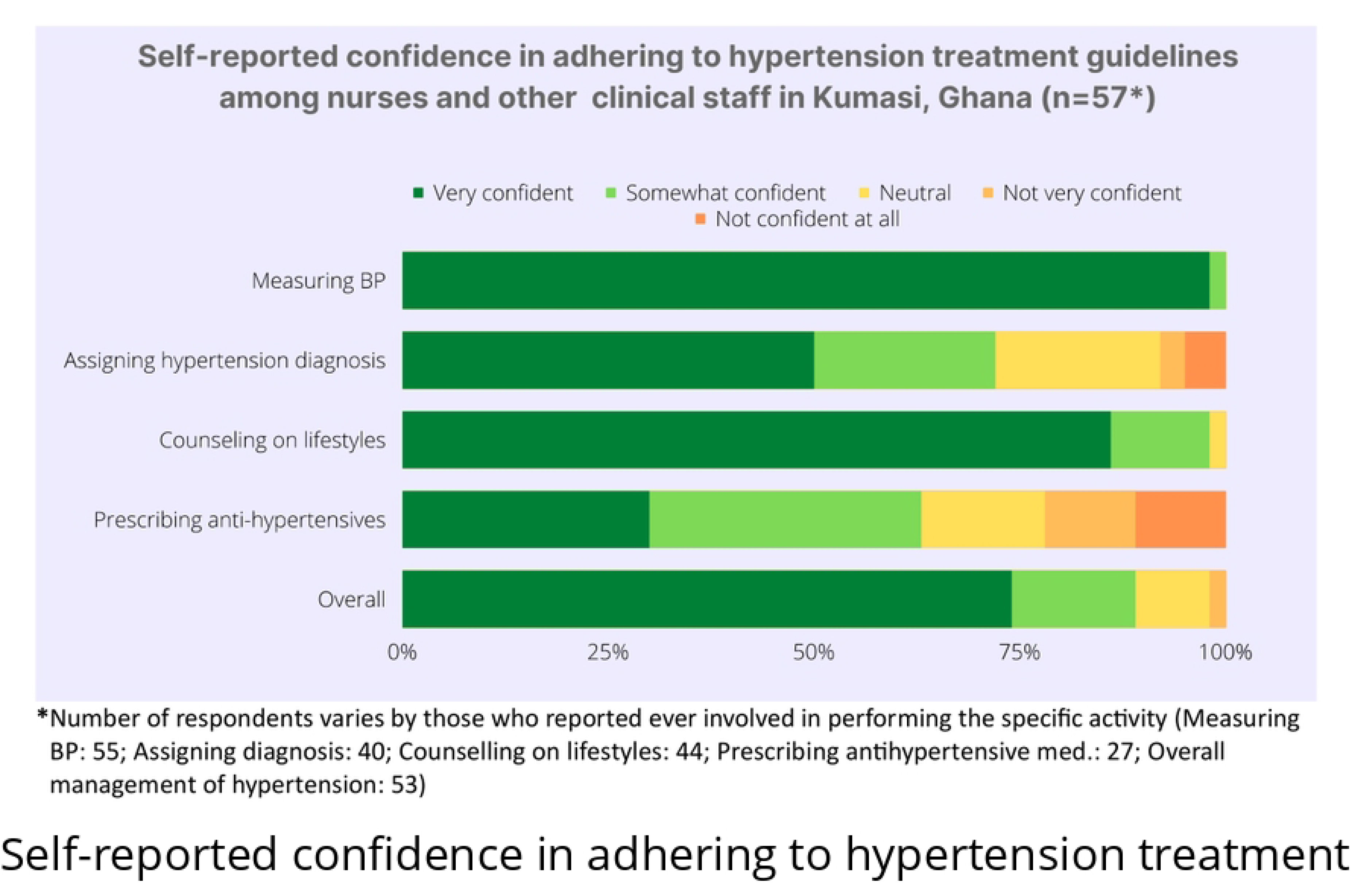
Self-reported confidence in adhering to hypertension treatment guidelines among nurses and other clinical staff (except medical doctors) in Kumasi, Ghana (n = 57)

#### Health facility level predictors of systolic and diastolic BP and BP control

At the health facility level, the facility ownership and patient-to-physician or physician assistant ratio predicted both systolic and diastolic BP and provider training showed association with diastolic BP (Tables 6).

Compared to private health facilities, receiving care at government health facilities was associated with a 19 mmHg (Coefficient: -19.4; 95% CI: -33.58, -5.22) and 13 mmHg (Coefficient: -13.48; 95% CI: -22.21, -4.76) lower systolic and diastolic BP, respectively, and receiving care at CHAG health facilities was associated with a 23 mmHg (Coefficient: -23.25; 95% CI: -39.98, -6.52) and 14 mmHg (Coefficient: -14.31; 95% CI: -24.6, -4.01) lower systolic and diastolic BP, respectively.

Receiving care at health facilities where a physician or physician assistant consulted 140 to 280 patients per month in OPD was associated with a 23 mmHg (Coefficient: 23.06 95% CI: 10.06, 36.05) and 15 mmHg (Coefficient: 15.52; 95% CI: 7.53, 23.52) higher systolic and diastolic BP, respectively, than where those clinicians consulted less than 140 patients per month in OPD. At facilities where a physician or physician assistant consulted 280 or more patients per month, there was a 27 mmHg (Coefficient: 23.06 95% CI: 10.06, 36.05) average higher systolic BP. Oddly, patients at health facilities that reported having at least one provider who received training in cardiovascular diseases in the last 2 years had a 7 mmHg higher diastolic BP (Coefficient: 6.8; 95% CI: 0.62, 12.98).

In the unadjusted and fully adjusted mixed-effects logistic regression models, patient-to-physician or physician assistant ratio (at the health facility level) predicted the patients’ BP control status (Table 6). Patients receiving care at health facilities where a physician or physician assistant consulted 140 to 280 patients per month were five times less likely (OR: 0.18; 95% CI: 0.05, 0.74) to have controlled BP than those who received care where a physician or physician assistant consulted less than 140 patients per month.

**Table 6.** Mixed-Effects Linear Regression of Health Facility Factors of Systolic and Diastolic Blood Pressure and Blood Pressure Control in Kumasi, Ghana (n = 221 patients, 15 health facilities)

|  | Adjusted SBP<br>Coefficients, 95% CI* | Adjusted DBP<br>Coefficients, 95% CI* | Adjusted BP<br>Control OR, 95%<br>CI* |
| --- | --- | --- | --- |
| **Facility level factors** |  |  |  |
| **Facility ownership** |  |  |  |
| *Private* | Ref. | Ref. | Ref. |
| *Government* | -19.4 (-33.58, -5.22) † | -13.48 (-22.21, -4.76) † | 2.2 (0.49, 9.96) |
| *Christian Health Association of Ghana* | -23.25 (-39.98, -6.52) † | -14.31 (-24.6, -4.01) † | 3.4 (0.58, 19.85) |
| **Hypertension Clinic** |  |  |  |
| *General OPD only* | Ref. | Ref. | Ref. |
| *Yes* | -2.27 (-9.08, 4.53) | -2.4 (-6.59, 1.79) | 1.26 (0.61, 2.58) |
| **Provider training** |  |  |  |
| *No* | Ref. | Ref. | Ref. |
| *Yes* | 6.4 (-3.65, 16.45) | 6.8 (0.62, 12.98) † | 0.38 (0.13, 1.10) |
| **Facility size by bed count** |  |  |  |
| *<50 beds* | Ref. | Ref. | Ref. |
| *50 to <100 beds* | 1.48 (-11.15, 14.12) | 0.81 (-6.96, 8.59) | 0.67 (0.18, 2.49) |
| *≥100 beds* | -4.06 (-17.75, 9.63) | -0.62 (-9.05, 7.8) | 1.54 (0.37, 6.51) |
| **All first line medications available** |  |  |  |
| *No* | Ref. | Ref. | Ref. |
| *Yes* | -5.25 (-17.51, 7.01) | -1.06 (-8.61, 6.48) | 1.6 (0.43, 5.89) |
| **Patient to nurse ratio (patients per month per nurse)** |  |  |  |
| *<20* | Ref. | Ref. | Ref. |
| *20 to <40* | -6.1 (-16.95, 4.76) | -5.35 (-12.03, 1.33) | 1.3 (0.42, 4.08) |
| *≥40* | -1.72 (-12.5, 9.06) | -3.34 (-9.98, 3.29) | 0.93 (0.3, 2.88) |
| **Patient to physician or physician assistant ratio (patients per month per clinician)** |  |  |  |
| *<140* | Ref. | Ref. | Ref. |
| *140 to <280* | 23.06 (10.06, 36.05) † | 15.52 (7.53, 23.52) † | 0.18 (0.05, 0.74) † |
| *≥280* | 27.24 (6.59, 47.88) † | 12.83 (0.12, 25.53) † | 0.27 (0.03, 2.38) |

\*Adjusted for all other variables plus age, sex, education, employment status; OPD: Outpatient Department; SBP: Systolic Blood Pressure; DBP: Diastolic Blood Pressure.
†Statistically significant (p<0.05)

## Discussion

The purpose of this study was to explore the health facility readiness for hypertension management and assess the patient-, provider-, and health facility-level factors of systolic and diastolic BP and BP control in Ghana. Most health facilities had at least one trained provider, had hypertension guidelines, BP measurement equipment, and basic laboratory exams. Conversely, the availability of patient educational materials and all first-line antihypertensive medications was sub-optimal. We found that nearly half of the patient participants had a controlled BP. Patients’ education, travel duration to the health facility, and adherence to treatment predicted BP outcomes. Providers had high knowledge of hypertension guidelines and reported being very confident in their adherence to those guidelines in practice. At health facility level, the facility ownership, the patient-to-physician or physician assistant ratio, and provider training predicted the patient BP outcomes.

Our findings are comparable to the findings of the 2019 May Measurement Month conducted in Kumasi, Ashanti Region, Ghana [29]. In that study, 49% of the patients on treatment for hypertension had a controlled BP [29]. Different studies, however, have reported slightly different levels of controlled BP among patients on hypertension treatment. The 2018 May Measurement Month conducted in Ashanti Region reported a 32% controlled BP among patients on hypertension treatment [30]. A different community-based study conducted in Bono Region reported a 41% BP control [31], and a study which recruited patients from two hospitals in greater Accra reported a 23% BP control level [32]. It is, however, necessary to understand that only 30% of patients aware of their hypertension status are initiated on treatment, bringing the overall estimate of people with hypertension with controlled BP down to 6% [3].

Patients with a secondary education had roughly 8 mmHg lower systolic BP than their counterparts with primary or lower level of education. These findings are similar to those of the previous literature. In a study conducted in Accra, patients with junior high and secondary education had 3 times the odds of controlled BP than patients with no formal education [32]. The level of education is one of the indicators of socio-determinants of health where higher education is often associated with higher socioeconomic status, social networks, and access to better quality of healthcare. Studies in different SSA countries demonstrate a relationship, often mediated by body mass index, between wealth and hypertension, where higher socioeconomic status is often associated with poor BP outcomes (higher BP) [33–35]. In the analysis of the 2014 Demographic and Health Survey data in Ghana, Amegah and Nayha (2018) demonstrated that higher educational attainment attenuated the relationship between wealth and BP [33]. In their analysis, affluent people with primary education had up to 8 times higher odds of elevated BP than their poorest counterparts while there was no association between wealth and BP status among people with a university degree [33]. However, poor and less educated people are at increased risk for poor BP outcomes, as demonstrated by a study conducted in eight SSA countries [36]. In that multinational study, individuals with low wealth index in both low- and middle-income countries (LMICs) had 88% higher odds of uncontrolled BP than their counterparts with higher wealth index [36]. Interventions for addressing hypertension will have to include specific measures for socioeconomically disadvantaged populations.

We found that travelling for an hour or more to the health facility for hypertension care was associated with roughly 7 mmHg higher diastolic BP. The distance of travel to the health facility is an important factor of health services accessibility and utilization [37]. The international standards specify that 80% of a given country’s population should have access to a hospital within 2 hours of walking distance for a timely management of emergencies. Yet, the literature lacks consistency as to which practical distance or travel time from the health center is associated with optimally good health outcomes. Some studies suggest 30 minutes from health centers, others 45 minutes, and others consider one hour as the cap for living in the served radius of a community health center [38–40]. The 2019 Ghana Living Standards Survey reported that on average 20% of people in Ghana travelled more than an hour to the nearest health facility [41], and another study reported that 14% of the population in Ghana travel more than two hours to any nearest public hospital [42]. One significant challenge with hypertension care in Ghana is the lack of emphasis on strengthening hypertension care services at the community-level health facilities [43]. Since there are no guidelines and enforcement of systems for managing or following up patients with hypertension at the low levels of the health systems (community-level health facilities), and systems such as mobile clinics or digital healthcare are absent, patients must travel long distances to hospitals for their regular hypertension follow-up appointments [43]. On the hospital side, the patient overload leads to quicker depletion of resources, longer waiting times, and the overall poor quality of services, hence poor health outcomes. In a similarly low-income SSA country, Rwanda, a study demonstrated that after removing the distance barriers of care access through the decentralization of hypertension care services to community health centers, there was no significant difference in achieving BP targets between patients followed up at the district hospitals versus community health centers [44]. Strategies for improving the geographical access of hypertension services are very much needed in Ghana.

In the current study, we found that higher hypertension treatment adherence scores were associated with lower diastolic BP. These findings corroborate the previous literature. In their study, Sarfo and colleagues (2020) showed that each one-unit increase in the scores of adherence to treatment was associated with a quarter unit decline in diastolic BP and a 3% higher odds of BP control [45]. Multiple factors at the patient-, provider-, and health facility level influence the patient adherence to treatment. At the patient-level, the forgetfulness, alcohol consumption and smoking behaviors, and the lack of motivation with the understanding that hypertension is a chronic condition requiring ongoing treatment are some of the reasons that hinder compliance to treatment [46,47]. The provider factors include the competence in educating patients on their health situation and the goal for treatment, and their own knowledge on hypertension could influence patients’ adherence to treatment. At the health facility and health systems level, the availability, accessibility and affordability of the medications have been expressed as major barriers to patients’ adherence to treatment [46]. The success of any strategy to manage hypertension will in many ways require finding measures for addressing factors of non-compliance with hypertension treatment.

Healthcare providers had high knowledge scores on hypertension guidelines and reported a high overall self-confidence in adhering to guidelines on tasks that were in their scope of practice.

While there is limited literature on healthcare providers’ objective knowledge of hypertension guidelines, extensive literature exists about healthcare providers’ perceptions on barriers to hypertension management in Ghana—alluding more to their lack of knowledge on hypertension and the need for more training [48,49]. A qualitative interview with healthcare providers at community health centers in the Upper East Region of Ghana reported that providers felt that they had inadequate knowledge on cardiovascular disease screening, treatment, and prescribing medications for cardiovascular diseases, and expressed the need for regular training sessions, workshops, and mentorship opportunities on cardiovascular diseases management [43]. In our findings, many health facilities reported that at least one healthcare provider had received training on hypertension management within two years, which could explain our findings of higher knowledge scores and confidence. Yet, the training did not translate to better BP outcomes. In our assessment, the most common type of training available for healthcare providers was the job orientation for new hires. We think that the low patients’ outcomes among facilities with trained providers could be explained by the new and unexperienced healthcare providers. Beyond provider training, however, there should be a system for flagging patients with uncontrolled BP to identify patient-specific support measures to help them better adhere to healthy behaviors and prescribed medications. More studies exploring the healthcare providers’ knowledge are needed to help strategize the needed content areas to cover while training healthcare providers in cardiovascular diseases management. Finally, the structured training accompanied with mentoring could be necessary to ensure that the acquired knowledge appropriately translated to practice, hence better patients outcomes.

We found that at facilities where a physician or physician assistant consulted 280 or more patients per month, patients had an average of 27-mmHg higher systolic BP and were five times less likely to have controlled BP. These findings speak to the shortage of healthcare providers, especially those in charge of doing consultation and prescribing medications for patients with hypertension (physicians, and physician assistants) and the associated poor BP outcomes. With a long queue of patients outside, the physician or physician assistant can only rush through the consultation, hence lacking adequate time to appropriately assess the patient needs to adjust the treatment accordingly. Like in many other LMICs, Ghana health policy still does not permit nurses to independently diagnose and prescribe antihypertensive medications [43,50]. Multiple studies have successfully trained nurses to diagnose hypertension and prescribe medications and patients have been very receptive of this approach [43,49,51,52]. Yet, practical and policy challenges continue to impede the implementation of task-shifting strategy. Amid the challenges of maintaining the high numbers of medical doctors and physicians, the next best approach will be for policymakers to refer to the existing evidence and quickly institute and reinforce a policy on training nurses to manage hypertension, including the diagnosis and prescription of antihypertensive medications.

We found that receiving care at private health facilities was associated with poor BP outcomes. These findings are counterintuitive, given that most of private health facilities are created because of inefficiencies in public health facilities and are often reported to have shorter wait times, better patient-provider relationships, and higher overall user satisfaction [53]. While it is not very clear why hypertension care in private health facilities is associated with poor outcomes, a few elements could offer some explanations. One strategy that has shown to be effective in hypertension management is team-based care—where healthcare professionals collaborate to manage patients with hypertension [54]. Yet, to minimize expenses, private health facilities de-emphasize the need to hire all key healthcare professionals such as pharmacists, dieticians, community health workers and others. Further, many private health facilities may not have the financial capacity to employ full-time cardiologists or medical doctors, while public health facilities may have them in addition to the other allied health professionals who are essential in team-based hypertension management.

Another key aspect of care in private versus public health facilities is the cost of care. The cost of care and the patient out-of-pocket spending are usually higher in private than government or quasi-governmental health facilities facilities [53]. The high cost of care makes it that poor people cannot attend private health facilities, but even the so-called wealthy might not be rich enough to afford the care and associated costs. Looking for ways to cut spending, patients could opt for poor utilization of health services or not purchasing some medications, hence leading to poor BP health outcomes. Like public health facilities, many private health facilities in Ghana provide services to patients insured by the National Health Insurance Scheme (NHIA). Yet, previous literature has highlighted challenges with the NHIA, including major delays in reimbursements that impede timely programming and health planning, especially in private health facilities, which rely solely on these funds for all facility activities [40,43]. These lead to poor access to medications and other key equipment and justification of higher out-of-pocket charges that impact patients’ outcomes.

Our study comes with some limitations. First, with the use of cross-sectional design, we could not longitudinally assess the change in BP or achievement of BP control and associated patient-, provider-, and facility-level factors. Second, our study was conducted at a small number of health facilities, which did not allow sufficient statistical power to detect small associations between facility-level predictors with the outcomes of interest. Third, in Ghana and many other countries in SSA, there are no systems for patients to maintain one primary healthcare provider, hence, we could not link patients’ outcomes with individual healthcare providers. We hence aggregated variables of provider-level characteristics, knowledge scores, and self-reported adherence to hypertension guidelines but could not link them directly to patients’ BP outcomes. Additionally, some assessments of the patient- and provider-level predictors were based on self-reported measures which could be associated with recall and social desirability bias. Last, the Service Availability and Readiness Assessment tool we used to measure health facility data has not been extensively utilized in other settings to measure hypertension service readiness, which limits comparison of findings across studies and settings. Despite the listed limitations, our study has major strengths in that it incorporates a broader viewpoint (from patient up to the facility-level) on the factors associated with hypertension service provision and patients’ BP outcomes. This study provides data that is vital to entities looking to explore the multi-level factors of hypertension care and outcomes at a broader scale or looking to implement interventions for hypertension management.

## CONCLUSION

In conclusion, hypertension control among patients with hypertension on treatment is low. Patients’ education and access to treatment, facility type of management, and provider density are associated with BP outcomes. Providers expressed high confidence in adhering to guidelines, but provider training was associated with poor outcomes, highlighting the lack of experience in hypertension management. Interventions targeting the patient-, provider-, and health system-levels will be needed to successfully address the hypertension burden. A large-scale study exploring the patient, provider, health facilities, and overall system factors of hypertension care and outcomes is needed. Specifically, larger studies should explore hypertension services accessibility, provider knowledge to guide curriculum development, nurse-to-patient ratio, and overall health facility hypertension readiness characteristics on patients’ BP outcomes

## Declarations

### Ethics Statement

This study was approved by the committee on human research, publication, and ethics at Kwame Nkrumah University of Science and Technology – School of Medicine and Dentistry (Ref: CHRPE/AP/021/22) and the Johns Hopkins Medicine Institutional Review Board (IRB00218586). After securing the ethics committee approvals, we secured signed memoranda of understanding with each participating health facility detailing the study procedures, timelines, and potential individuals to be invited in the study. All patients and healthcare providers signed informed paper and electronic consent, respectively, prior to participating in the study.

## Data Availability

Dataset for the data presented in this manuscript is available by request to the corresponding author. Email Samuel Byiringiro at.

## Acknowledgment

We thank all the health facilities in Kumasi Ghana, healthcare providers, and patients who participated in the ADHINCRA study.

## Supporting information

S1 Table. STROBE Checklist

## References

1. Guwatudde D, Nankya-Mutyoba J, Kalyesubula R, Laurence C, Adebamowo C, Ajayi IO, et al. The burden of hypertension in sub-Saharan Africa: A four-country cross sectional study. BMC Public Health. 2015;15: 1211. doi:10.1186/s12889-015-2546-z

2. World Health Organization (WHO). Hypertension Fact Sheet. 2019 [cited 15 Apr 2020]. Available: https://www.who.int/news-room/fact-sheets/detail/hypertension

3. Bosu WK, Bosu DK. Prevalence, awareness and control of hypertension in Ghana: A systematic review and meta-analysis. PLoS ONE. 2021. doi:10.1371/journal.pone.0248137

4. Nyaaba GN, Stronks K, de-Graft Aikins A, Kengne AP, Agyemang C. Tracing Africa’s progress towards implementing the Non-Communicable Diseases Global action plan 2013-2020: a synthesis of WHO country profile reports. BMC Public Health. 2017;17: 297. doi:10.1186/s12889-017-4199-6

5. Iwelunmor J, Plange-Rhule J, Airhihenbuwa CO, Ezepue C, Ogedegbe O. A Narrative Synthesis of the Health Systems Factors Influencing Optimal Hypertension Control in Sub-Saharan Africa. PLoS One. 2015;10: e0130193–e0130193. doi:10.1371/journal.pone.0130193

6. Nulu S, Aronow WS, Frishman WH. Hypertension in Sub-Saharan Africa: A Contextual View of Patterns of Disease, Best Management, and Systems Issues. Cardiol Rev. 2016;24: 30–40. doi:10.1097/CRD.0000000000000083

7. Whelton PK, Carey RM, Aronow WS, Casey DE, Collins KJ, Himmelfarb CD, et al. 2017 ACC/AHA/AAPA/ABC/ACPM/AGS/APhA/ASH/ASPC/NMA/PCNA Guideline for the Prevention, Detection, Evaluation, and Management of High Blood Pressure in Adults. J Am Coll Cardiol. 2018;71: e127 LP-e248. doi:10.1016/j.jacc.2017.11.006

8. James PA, Oparil S, Carter BL, Cushman WC, Dennison-Himmelfarb C, Handler J, et al. 2014 Evidence-Based Guideline for the Management of High Blood Pressure in Adults: Report From the Panel Members Appointed to the Eighth Joint National Committee (JNC 8). JAMA. 2014;311: 507–520. doi:10.1001/jama.2013.284427

9. Unger T, Borghi C, Charchar F, Khan NA, Poulter NR, Prabhakaran D, et al. 2020 International Society of Hypertension Global Hypertension Practice Guidelines. Hypertension. 2020;75: 1334–1357. doi:10.1161/HYPERTENSIONAHA.120.15026

10. Jeemon P, Séverin T, Amodeo C, Balabanova D, Campbell NRC, Gaita D, et al. World heart federation roadmap for hypertension – A 2021 update. Glob Heart. 2021;16: 63. doi:10.5334/GH.1066/METRICS/

11. Suzuki Y, Kaneko H, Yano Y, Okada A, Matsuoka S, Fujiu K, et al. Reduction in blood pressure for people with isolated diastolic hypertension and cardiovascular outcomes. Eur J Prev Cardiol. 2022 [cited 10 Jan 2023]. doi:10.1093/EURJPC/ZWAC278

12. Flint AC, Conell C, Ren X, Banki NM, Chan SL, Rao VA, et al. Effect of Systolic and Diastolic Blood Pressure on Cardiovascular Outcomes. New England Journal of Medicine. 2019;381: 243–251. doi:10.1056/NEJMOA1803180/SUPPL_FILE/NEJMOA1803180_DISCLOSURES.PDF

13. Chaix B, Bean K, Leal C, Thomas F, Havard S, Evans D, et al. Individual/neighborhood social factors and blood pressure in the RECORD Cohort Study: which risk factors explain the associations? Hypertension. 2010;55: 769–75. doi:10.1161/HYPERTENSIONAHA.109.143206

14. Wagner EH. Chronic disease management: what will it take to improve care for chronic illness? Effective clinical practice : ECP. United States; 1998. pp. 2–4.

15. Owusu MF, Basu A, Barnett P. Hypertension and diabetes management: a policy perspective from Ghana. J Health Organ Manag. 2018/11/09. 2019;33: 35–50. doi:10.1108/JHOM-03-2018-0076

16. World Health Organization (WHO). Monitoring the Building Blocks of Health Systems: A handbook of indicators and their meaurement strategies. World Health Organization. Geneva; 2010. doi:10.1146/annurev.ecolsys.35.021103.105711

17. Ahafo B, Accra G, East U, West U, Executives C, Comity T, et al. Ghana Districts - A repository of all districts in the republic of Ghana. 2012; 2–3.

18. Ghana Statistical Service. Population by Regions. 2019 [cited 18 Nov 2020]. doi:10.1128/AAC.03728-14

19. Ghana Statistical Service. Kumasi metropolitan. 2010; 92.

20. Ghana Open data initiative. Health Facilities | Ghana Open Data Initiative. 2016 [cited 16 Jun 2020]. Available: https://data.gov.gh/dataset/health-facilities-ghana/resource/aa5b8e2f-a2e7-4aa6-9fef-cc31e1d1aa90#%7Bquery:%7Bfilters:[%7Btype:!term,field:!Region,term:!ashanti%7D]%7D,view-graph:%7BgraphOptions:%7Bhooks:%7BprocessOffset:%7B%7D,bindEvents:%7B%7D%7D%7D%7D,

21. Cuschieri S. The STROBE guidelines. Saudi J Anaesth. 2019;13: S31. doi:10.4103/SJA.SJA_543_18

22. Muntner P, Shimbo D, Carey RM, Charleston JB, Gaillard T, Misra S, et al. Measurement of Blood Pressure in Humans: A Scientific Statement From the American Heart Association. Hypertension. 2019;73: E35–E66. doi:10.1161/HYP.0000000000000087

23. NATIONAL GUIDELINES FOR THE MANAGEMENT OF CARDIOVASCULAR DISEASES First Edition. Accra; 2019.

24. Kim MT, Hill MN, Bone LR, Levine DM. Development and testing of the Hill-Bone Compliance to High Blood Pressure Therapy Scale. Prog Cardiovasc Nurs. 2000;15: 90–96. doi:10.1111/J.1751-7117.2000.TB00211.X

25. Baldridge A, Orji A, Omitiran K, Guo M, Ajisegiri S, Ojo T, et al. Capacity and Site Readiness for Hypertension Control Program Implementation in the Federal Capital Territory of Nigeria: A Cross-Sectional Study. Health Serv Res. 2020;55: 67–67. doi:10.1111/1475-6773.13421

26. National Standard Treatment Guidelines 7th Edition. Accra; 2017. Available: https://www.moh.gov.gh/wp-content/uploads/2020/07/GHANA-STG-2017-1.pdf

27. Ministry of Health Ghana National Drugs Programme (GNDP) - Essential Medicines List 7th Edition. Accra; 2017. Available: https://www.moh.gov.gh/wp-content/uploads/2020/07/GHANA-EML-2017.pdf

28. Sarfo F, Treiber F, Gebregziabher M, Adamu S, Patel S, Nichols M, et al. PINGS (Phone- Based Intervention Under Nurse Guidance After Stroke): Interim Results of a Pilot Randomized Controlled Trial. Stroke. 2017/12/08. 2018;49: 236–239. doi:10.1161/STROKEAHA.117.019591

29. Tannor EK, Nyarko OO, Adu-Boakye Y, Owusu Konadu S, Opoku G, Ankobea-Kokroe F, et al. Prevalence of Hypertension in Ghana: Analysis of an Awareness and Screening Campaign in 2019. https://doi.org/101177/11795468221120092. 2022;16. doi:10.1177/11795468221120092

30. Tannor EK, Nyarko OO, Adu-Boakye Y, Owusu Konadu S, Opoku G, Ankobea-Kokroe F, et al. Burden of hypertension in Ghana – Analysis of awareness and screening campaign in the Ashanti Region of Ghana. JRSM Cardiovasc Dis. 2022;11: 204800402210755. doi:10.1177/20480040221075521

31. Dosoo DK, Nyame S, Enuameh Y, Ayetey H, Danwonno H, Twumasi M, et al. Prevalence of Hypertension in the Middle Belt of Ghana: A Community-Based Screening Study. Int J Hypertens. 2019;2019. doi:10.1155/2019/1089578

32. Okai DE, Manu A, Amoah EM, Laar A, Akamah J, Torpey K. Patient-level factors influencing hypertension control in adults in Accra, Ghana. BMC Cardiovasc Disord. 2020;20. doi:10.1186/S12872-020-01370-Y

33. Amegah AK, Näyhä S. Educational attainment modifies the association of wealth status with elevated blood pressure in the Ghanaian population. Heliyon. 2018;4: 711. doi:10.1016/J.HELIYON.2018.E00711

34. Banchani E, Tenkorang EY, Midodzi W. Examining the effects of individual and neighbourhood socioeconomic status/wealth on hypertension among women in the Greater Accra Region of Ghana. Health Soc Care Community. 2022;30: 714–725. doi:10.1111/HSC.13185

35. Mustapha A, Ssekasanvu J, Chen I, Grabowski MK, Ssekubugu R, Kigozi G, et al. Hypertension and Socioeconomic Status in South Central Uganda: A Population-Based Cohort Study. Glob Heart. 2022;17. doi:10.5334/GH.1088/METRICS/

36. Antignac M, Diop IB, de Terline DMQ, Kramoh KE, Balde DM, Dzudie A, et al. Socioeconomic status and hypertension control in sub-saharan Africa the multination EIGHT study (evaluation of hypertension in Sub-Saharan Africa). Hypertension. 2018;71: 577–584. doi:10.1161/HYPERTENSIONAHA.117.10512

37. Sarkodie AO. The effect of the price of time on healthcare provider choice in Ghana. Humanities and Social Sciences Communications 2022 9:1. 2022;9: 1–7. doi:10.1057/s41599-022-01282-6

38. dos Anjos Luis A, Cabral P. Geographic accessibility to primary healthcare centers in Mozambique. Int J Equity Health. 2016;15: 1–13. doi:10.1186/S12939-016-0455-0

39. Stock R. Distance and the utilization of health facilities in rural Nigeria. Soc Sci Med. 1983;17: 563–570. doi:10.1016/0277-9536(83)90298-8

40. Koduah A, Nonvignon J, Colson A, Kurdi A, Morton A, van der Meer R, et al. Health systems, population and patient challenges for achieving universal health coverage for hypertension in Ghana. Health Policy Plan. 2021;36. doi:10.1093/heapol/czab088

41. Ghana Statistical Service. Ghana Living Standards Survey (GLSS). Accra; 2019 Jun.

42. Ouma PO, Maina J, Thuranira PN, Macharia PM, Alegana VA, English M, et al. Access to emergency hospital care provided by the public sector in sub-Saharan Africa in 2015: a geocoded inventory and spatial analysis. Lancet Glob Health. 2018;6: e342–e350. doi:10.1016/S2214-109X(17)30488-6

43. Laar AK, Adler AJ, Kotoh AM, Legido-Quigley H, Lange IL, Perel P, et al. Health system challenges to hypertension and related non-communicable diseases prevention and treatment: Perspectives from Ghanaian stakeholders. BMC Health Serv Res. 2019;19: 1–13. doi:10.1186/S12913-019-4571-6/TABLES/1

44. Ngoga G, Park PH, Borg R, Bukhman G, Ali E, Munyaneza F, et al. Outcomes of decentralizing hypertension care from district hospitals to health centers in Rwanda, 2013–2014. Public Health Action. 2019;9: 142. doi:10.5588/PHA.19.0007

45. Sarfo FS, Mobula L, Plange-Rhule J, Gebregziabher M, Ansong D, Sarfo-Kantanka O, et al. Longitudinal control of blood pressure among a cohort of Ghanaians with hypertension: A multicenter, hospital-based study. The Journal of Clinical Hypertension. 2020;22: 949. doi:10.1111/JCH.13873

46. Woode E, Boakye-Gyasi E, Obirikorang Y, Adu EA, Obirikorang C, Acheampong E, et al. Predictors of medication nonadherence among hypertensive clients in a Ghanaian population: Application of the Hill-Bone and Perceived Barriers to Treatment Compliance Scale. Health Sci Rep. 2022;5: e584. doi:10.1002/HSR2.584

47. Sarkodie E, Afriyie DK, Hutton-Nyameaye A, Amponsah SK. Adherence to drug therapy among hypertensive patients attending two district hospitals in Ghana. Afr Health Sci. 2020;20: 1355. doi:10.4314/AHS.V20I3.42

48. De-Graft Aikins A, Kushitor M, Koram K, Gyamfi S, Ogedegbe G. Chronic non-communicable diseases and the challenge of universal health coverage: Insights from community-based cardiovascular disease research in urban poor communities in Accra, Ghana. BMC Public Health. 2014;14: 1–9. doi:10.1186/1471-2458-14-S2-S3/TABLES/2

49. Iwelunmor J, Gyamfi J, Plange-Rhule J, Blackstone S, Quakyi NK, Ntim M, et al. Exploring stakeholders’ perceptions of a task-shifting strategy for hypertension control in Ghana: A qualitative study. BMC Public Health. 2017;17: 1–9. doi:10.1186/S12889-017-4127-9/PEER-REVIEW

50. Ogedegbe G, Gyamfi J, Plange-Rhule J, Surkis A, Rosenthal DM, Airhihenbuwa C, et al. Task shifting interventions for cardiovascular risk reduction in low-income and middle-income countries: a systematic review of randomised controlled trials. BMJ Open. 2014;4. doi:10.1136/BMJOPEN-2014-005983

51. Iwelunmor J, Gyamfi J, Plange-Rhule J, Blackstone S, Quakyi NK, Ntim M, et al. Exploring stakeholders’ perceptions of a task-shifting strategy for hypertension control in Ghana: a qualitative study. BMC Public Health. 2017;17: 216. doi:10.1186/s12889-017-4127-9

52. Iwelunmor J, Onakomaiya D, Gyamfi J, Nyame S, Apusiga K, Adjei K, et al. Adopting Task-Shifting Strategies for Hypertension Control in Ghana: Insights From a Realist Synthesis of Stakeholder Perceptions. Glob Heart. 2019;14: 119. doi:10.1016/j.gheart.2019.05.007

53. Tenkorang EY. Health Provider Characteristics and Choice of Health Care Facility among Ghanaian Health Seekers. . 2016;2: 160–170. doi:10.1080/23288604.2016.1171282

54. Ware LJ, Schutte AE. Team-based care for hypertensive patients is essential in low- and middle-income countries. The Journal of Clinical Hypertension. 2019;21: 1210–1211. doi:10.1111/JCH.13586

